# A Brick to a Bundle: Does Xylazine Paradoxically Contribute to Treatment-seeking and Reduced Fentanyl Use?

**DOI:** 10.1101/2025.04.11.25325471

**Authors:** Adams L. Sibley, Colin W. Miller, Elizabeth Joniak-Grant, Alice Bell, Malcolm Visnich, Steve Alsum, Nabarun Dasgupta

## Abstract

**Background:** Xylazine is a veterinary tranquilizer found in the unregulated drug supply in the United States. It appears alone or as an adulterant in fentanyl (“tranq dope”). Xylazine’s symptomatology is well described and includes skin and soft tissue damage, bradycardia, and loss of consciousness. However, little is known about whether and how substance use behaviors have changed as xylazine’s presence in street drugs has grown.

**Methods:** We conducted semi-structured in-depth interviews with people with recent overdose reversal experiences in two mid-sized midwestern cities (*n=*52). Interviews were part of a larger study on naloxone administration behaviors. Participants were asked about their knowledge and perceptions of local drug supply trends. Transcript data were analyzed using the rigorous and accelerated data reduction technique.

**Results:** Participants preferred fentanyl and heroin without xylazine. Most participants discussed adjusting opioid use toward safer practices: using less in amount or frequency, abstaining or seeking treatment, alternating use (e.g., ingesting xylazine only at night), or changing route of administration from injecting to smoking, snorting, or boofing (ingesting anally). Motivations for changes in use included not experiencing intended opioid agonist effects, fear of physical health risks, loss of functionality and productivity, and overdose concerns.

**Conclusion:** Findings suggest that xylazine is encouraging reduced fentanyl and heroin use. Our results corroborate laboratory, clinical, and behavioral studies showing that xylazine, which causes severe health harms, may also, paradoxically, be protective against fatal overdose. More research is needed on this phenomenon in light of recent downward trends in overdose mortality.

## 1. Introduction

Xylazine, a veterinary tranquilizer not approved for human use, has been routinely described in the United States’ unregulated drug supply. Xylazine has agonist properties at alpha-2 adrenergic, kappa opioid, dopamine, and sigma receptors (Bedard et al., 2024). It typically appears mixed with fentanyl or heroin, in a combination known as ‘tranq dope.’

Xylazine emerged as a heroin adulterant in Puerto Rico in the early 2000s, reaching the mainland (notably Philadelphia) by the end of the decade (Rodríguez et al., 2008; Wong et al., 2008). Its unexplained proliferation across the country exhibits an east-to-west trend, appearing earlier and with greater prevalence in mid-Atlantic states (Cano et al., 2024).

People who use drugs (Carroll, 2024) and medical professionals (Edinoff et al., 2024) report complications from xylazine exposure including bradycardia, hypotension, loss of consciousness, and severe skin and soft tissue damage that is long-lasting and often necrotic. Xylazine is undesirable to most people who use opioids (Hochheimer et al., 2024), but it is unclear whether and how its presence changes consumption behaviors. The purpose of this qualitative study, therefore, was to understand recent patterns of fentanyl and heroin use from the perspectives of individuals exposed to xylazine.

## 2. Methods

Data were collected as part of a larger mixed methods study on community-based naloxone utilization, including quantitative analysis of reversal data and analysis of drug samples collected by partner harm reduction programs in two mid-sized US cities in Michigan and Pennsylvania.

Xylazine was entrenched in the drug supply at both sites; [RESEARCH INSTITUTION] provides mail-in analysis services using gas chromatography-mass spectrometry (GCMS) (Wagner et al., 2023). Of 591 fentanyl samples analyzed (April 2023 to November 2024), the unadjusted presence of xylazine was 48.6% (*n*=222/457) at the Michigan site and 74.6% (*n*=100/134) at the Pennsylvania site.

Participants (*N*=52) in the present study were recruited from the partner harm reduction programs or by peer referral in Pennsylvania (*n=*27) and Michigan (*n*=25). Inclusion criteria: 1) aged 18 years or older, 2) administered naloxone in past 12 months, 3) currently using drugs with overdose risk or have direct social contacts who use drugs.

We conducted semi-structured in-depth interviews in October-November 2024, using an interview guide developed in partnership with epidemiologists and behavioral scientists from the Food and Drug Administration. Topics included: experiences reversing overdoses, attitudes toward overdose reversal agents, and perceptions of the current unregulated drug supply, including xylazine. Interviews were conducted at program sites, averaged 45 minutes, and were incentivized with $50 cash. The study was approved by the [RESEARCH INSTITUTION] Institutional Review Board (#24-0265). Participants provided verbal consent; no signatures or demographic information were collected to enhance confidentiality. People with lived experience were involved in the study design and collection, analysis, and reporting of the data.

Data were thematically analyzed using the RADaR (rigorous and accelerated data reduction) approach (Watkins, 2017) with three phases of data reduction, excerpt-level open coding, and collaborative interpretation (Appendix A). Below, we report new patterns of use in the wake of xylazine and motivations for these changes. Additional exemplifying excerpts are in Appendix B.

## 3. Results

Having learned about xylazine through news, social media, harm reduction and treatment programs, and social networks, participants were largely familiar with its effects and presence in unregulated opioid markets. Many experienced xylazine’s effects before putting a name to it (“Before I knew what it was, I was already seeing the effects, mostly just in abscesses that were happening”). No participants shared positive perceptions of xylazine (though there were limited secondhand accounts of preference), and most lamented its ubiquity in the supply (“You can’t find anything without it”). Participants overwhelmingly related that the presence of xylazine had driven their fentanyl consumption lower. (There were only four exceptions: two participants reported using more in recent months, one due to increased tolerance to fentanyl, and the other due to perceived shorter duration or “legs” of tranq dope; and two reported no change in use.) Otherwise, we categorized four predominant consumption patterns in the wake of xylazine:

1. using less in amount or frequency;
2. quitting or aspiring to recovery;
3. alternating use;
4. changing route of administration.

### 3.1 Changes in Use

#### 3.1.1 Using Less

Most participants who reported modifying opioid consumption discussed using less frequently (“At least every two hours” to “three times a day”), in smaller amounts (“You only need like a half of a stamp bag”), or with periods of temporary abstinence:

> I’m to the point where I’m like, in 30 years, I have not been able to quit doing dope. But I have days where I don’t do it. And that’s never happened. Never happened out of rehab, for 30 years. I’ve been a hardcore addict for about 30 years. And xylazine scares the fuck out of me.

For participants reporting amount, reductions were substantial: Several in Pennsylvania quantified their use as shrinking from about a brick (#50) to a bundle (#10) of 0.1g bags daily.

#### 3.1.2 Quitting or Aspiring to Recovery

Several participants reported stopping overall opioid use because of xylazine (“[Xylazine] scared me to the point where I knew I had to get clean”), often with aid of medications for opioid use disorder (“[Xylazine’s] what made me get on Subs [buprenorphine]”). Others reported a newfound desire to discontinue use (“It’s been getting to the point where, if everything is going to have so much xylazine in it, it’s making us want to just not use at all”).

#### 3.1.3 Alternating Use

A few participants continued use but alternated between xylazine-positive and xylazine-negative bags – for instance, using adulterated bags only to induce sleep at night, or alternating to alleviate ostensible xylazine withdrawal symptoms (“I feel sick now from…and I want the ones *with* xylazine. So, how I would do it, would be like, one with xylazine and one without xylazine, you know? I would, like, alternate”).

#### 3.1.4 Route of Administration

Finally, participants discussed recent transitions from injecting to smoking, snorting, or boofing (i.e., ingesting anally). Some believed that alternative routes allow for easier dose titration to avoid sedation, reduce risk of lesions, or prevent injection complications (“It ruins your shots. Like, once you get little blood in there, it just coagulates and you can’t even get it. Just waste it”).

### 3.2 Motivation for Changes in Consumption

Participants shared multiple reasons for reducing, stopping, or altering use. Generally, xylazine elicited emotions ranging from intense fear to exasperation, with some reaching a breaking point after years of unfulfilling use:

> There’s a lot of things that you will accept, if it was going to be as good as it was when you first started. You’ll go through a lot of bullshit to get there, but if it’s not even going to be that, then you’re kind of done.

We categorized four reasons for the observed consumption pattern explanations:

1. not experiencing intended effects;
2. physical health risks;
3. functional impacts;
4. overdose beliefs.

#### 3.2.1 Not Experiencing Intended Effects

Participants bemoaned no longer feeling the desired psychoactive effects of opioids (“When you do heroin, it feels good. Obviously. That’s why we do it. But that shit [xylazine], it doesn’t”). Specific expected sensations missing with tranq dope were euphoria, pain relief, and emotional numbing. More immediately, participants reported tranq dope failed to curb opioid withdrawal symptoms (“It’s not even making me right anymore”).

#### 3.2.2 Physical Health Risks

Skin and soft tissue damage commonly motivated reduced use (“Breakfast, lunch, and dinner, I try to do three shots a day and just leave it at that because I don’t like having abscesses all over […] It makes me not want to leave my house”). Participants also expressed fear of limb loss:

> The biggest reason was my wife got a couple of the little wounds on her. […] And, I’m like, if […] she has to get her leg amputated or her arm amputated, like, cause it’s— I’m the one who fucked up.

Additional health risks participants hoped to avoid that were presumed to be due to xylazine included hallucinations and injection site complications (“Typically if you missed [a vein], it would get itchy and, like, red and bumpy. This one [xylazine], as soon as you missed, it was like lightning under your skin”).

#### 3.2.3 Functional Impacts

Xylazine can induce hours of unconsciousness or a “zombie-like” state to the frustration, embarrassment, or bewilderment of many participants (“I even taped myself because I had to see how this was happening”). Unintended sedation hindered many from achieving an expected level of functionality and productivity (“I use a lot less, that’s for sure. If you do too much of it, you’re just going straight to sleep. You waste the whole day”).

#### 3.2.4 Overdose Beliefs

Some participants contended tranq dope was “stronger,” potentiating overdose risk (“I’ve almost died off one bag, and I’m a veteran with this shit”), or made overdoses more difficult to reverse with naloxone (“They don’t pop out of it like a Pop-Tart anymore”). The consequent fear of mortality drove some participants to reduce or discontinue use:

> I’ve done an awful lot of reversals and I know [xylazine] affects your central nervous system and keeps you—Narcan doesn’t help reviving the respiratory part of it. So I had began using less and less until I had an intervention, essentially.

## 4. Discussion

In this qualitative study, we explored changes in drug consumption behaviors in the wake of xylazine’s appearance in the unregulated drug supply. With few exceptions, participants described recent attempts at safer use – using less, changing route of administration, or abstaining entirely. Participants with decades of experience recalled adapting to previous supply transitions (“We got used to [fentanyl] a long time ago, still not what we’d prefer over actual heroin, but it is what it is”). Xylazine, however, represented a novel inflection point, with some resigned that the current supply is no longer worth the pleasures of use, nor the pains of dependence.

While the human health risks of xylazine consumption are well-documented, our findings indicate that xylazine in the unregulated drug supply may have the unexpected effect of reducing fentanyl consumption. Findings are consistent elsewhere. Reduced self-administration has been documented in rodent models, where xylazine coadministration suppresses fentanyl consumption (Bedard et al., 2024; Khatri et al., 2024; Sadek et al., 2024); observational clinical research shows xylazine may be protective against cardiac arrest, oxygen distress, and fatality in fentanyl poisonings (Hays et al., 2024; Love et al., 2023); and similar shifts in perception and behavior have been discussed by Reddit users (Heidari et al., 2024). Collectively, these lines of evidence start to build the case for a paradoxical protective-against-fatal-overdose effect of xylazine along multiple causal pathways.

A key area of future investigation pertains to how xylazine-related behavioral shifts may influence population-level overdose mortality. In mid-2023, overdose deaths in the United States started decreasing, with annualized mortality dropping 30% from 2023-2024 in some states (Dyer, 2024). The east-to-west pattern of overdose declines parallels the spread of xylazine in the unregulated drug supply (Dyer, 2024).

However, the declines in overdose rates are surely more complex than the emergence of a single adulterant (Dyer, 2024). We can only conclude from our study that xylazine may have changed people’s use of fentanyl in the sites we investigated, often in compensatory ways to decrease risk of severe skin wounds and overdose fatality. If indeed xylazine is contributing to safer consumption behaviors (Heidari et al., 2024), then harm reduction interventions focused on safer use should be redoubled while risk salience is high and supply satisfaction remains at its nadir.

From a policy perspective, official measures (Sugarman et al., 2024) restricting xylazine should be evaluated in light of affecting overdose mortality. More broadly, international Drug Control Conventions have limited antecedents for evaluating an adulterant that simultaneously causes one kind of severe harm (skin wounds), and yet may have protective effect on another (fentanyl overdose). We urge caution against reactive policy measures that interrupt the unregulated supply of alpha-2 adrenergic agonists (xylazine and (dex)medetomidine): Our data suggest that the abrupt disappearance of these additives has the potential to reverse the observed behavioral changes described above, and lead to a rebound in more street fentanyl consumption.

In addition to restricted generalizability, a limitation of this study is that we cannot explain why illicit drug producers would want clients to reduce consumption. Nor can we explicate the sustained proliferation of an additive that customers strongly view as undesirable, though there are other ongoing examples (Shover et al., 2024). The presence of xylazine in the unregulated drug supply does not conform to our understanding of previous drug outbreaks, requiring more nuanced investigation.

## Data Availability

All data produced in the present study are available upon reasonable request to the authors

## Acknowledgements

The authors wish to thank Jana McAninch, Blair Coleman, and the rest of the FDA project advisory group for feedback on this manuscript.

### Appendix A: Qualitative Analysis

Interviews were audio-recorded then transcribed with the *whisper* package in Python 3.9 then manually corrected by ALS, who performed a close reading and memoing of the interview data. The analysis team (ALS, CWM, ND) then met to discuss the data, noting salience of xylazine to drug consumption behaviors among participants and setting the research question:

> How have participant drug consumption behaviors shifted in light of xylazine, and what factors may explain these shifts?

Data were analyzed using the RADaR (rigorous and accelerated data reduction) approach. Transcript excerpts were imported into an all-inclusive data table in Microsoft Excel then reduced (i.e., shortening or removing excerpts) and analyzed across three phases. In phase one, we memoed on potential themes and divided the data into separate tables by topic (changes in use, feelings toward xylazine, learning about xylazine). In phase two, we open-coded the excerpts in each table and reduced the data further. In phase three, we refined our open codes into a final (mixed inductive-deductive) thematic codebook which were then applied to each table. Excerpts were collated into code reports and interpreted by the analysis team to determine initial themes; code reports and thematic conceptual models were then reviewed by remaining team members and themes were finalized.

### Appendix B: Sample of Exemplary Excerpts by Theme

#### Changes in Use

##### Using Less

Using a lot less, I think. I don’t use as often I guess is part of it. But another thing is it’s really scary with the xylazine, I even taped myself because I had to see how this was happening. So I just pass out and I just wake up just you know what I mean. There’s no nod, there’s no nod with this thing— (I: You just go straight to—) Yeah. Yeah, it’s just straight out. Wake up a couple hours later. It’s like. Yeah, so and I use by myself so it was kind of scary there for a minute. (I: Oh, so you taped yourself to see what it looked like –) Yeah, yeah. (I: And you just kind of passed out.) Yeah, because I was, I’d just wake up like I don’t, I didn’t remember, there wasn’t even a nod, like I just wake up all of a sudden, I don’t remember anything, I remember taking a shot and everything, but I don’t remember… And then yeah, so I finally, I taped myself because I had to see you know how fast I’m going out and it was just like instant dude, like.

Probably just, yeah, just how much I use. Yeah, yeah. For me generally like you try, you generally use a little bit in the morning and my, like getting high a little more at night after work, so that’s it. Yeah, yeah, nothing, not usually during the day I’m at work, but then first thing in the morning and last thing at night, so yeah.

Yeah, it made me a lot more concerned with things. I had started intentionally using less and less. I’ve done an awful lot of reversals and I know that particular one also affects your central nervous system and keeps you, Narcan doesn’t help reviving the respiratory part of it. So I had began using less and less until I had an intervention, essentially. I had a warrant I was running from for a while and it caught up to me, it was like two years ago. I went to a county jail and ended up detoxing in there. A whole lot of my overdose reversals have been from people from a period of abstinence or when they get out of jail, mostly when people get out of jail and go out. What is it? After two days, your tolerance is reduced by 50 percent. Just that alone. But the majority of the time, I couldn’t believe how little it would take for somebody to use again to go blue in the lips and quit breathing and need Narcan.

Definitely how much because I was scared. (I: Scared? What did you do different?) I would just do less. I don’t know, because normally I’ve been getting high for like 15 years and I have a huge tolerance from like a crazy amount. So I was doing like a gram just to get unsick. Taking it all in at once so then I would just, I lowered it down to like a quarter, almost to a half a gram.

OK. So I’d say in the last six months around [city]… But as far as xylazine, it hit here around six months ago. It started hitting hard. In my experience, it’s tranq. It puts you out. I’ve seen some of the worst wounds that I’ve ever seen in my life. I’ve seen wounds that, like my girlfriend, she has one, it just doesn’t heal. She thinks it does. And then it just busts back open, and there’s black tissue under there, black muscle. And yeah, it hasn’t been good, man. It hasn’t been a good experience Yeah, xylazine, it’s just nasty shit. I’ve been an addict for 30 years. If it’s got xylazine, I don’t want it. I don’t want to do that. But this, it’s just everywhere now. I’m to the point where I’m like, in 30 years, I have not been able to quit doing dope. But I have days where I don’t do it. And that’s never happened. Never happened out of rehab, for 30 years. I’ve been a hard-core addict for about 30 years. And xylazine scares the fuck out of me. I don’t want to lose limbs. I don’t want to see people lose limbs. And it’s just nasty shit.

I’m way more careful and I use like a lot less. I try to use like, I kind of try to do, I know it sounds dumb, but like breakfast, lunch, and dinner, I try to do one, three shots a day and just leave it at that because I don’t like having abscesses all over and stuff like that. It makes me not want to leave my house and so I’ll be at home more often. Yeah. (I: So you use breakfast, lunch, and dinner. How much would you, how often would you use before that?) Um, I’d probably, I don’t know, probably at least every two hours. Every three hours. (I: Mm-hmm. So you’re using less frequently since xylazine showed up?) Yeah. (I: And that’s mostly because of the, you know, abscesses and the other physical effects?) Yeah. (I: Okay. Right. Um.) Yeah, it makes me very sleepy.

Uh, it decreased. Using a lot less. I’d probably go from a brick a day to probably about a bundle. To maybe a bundle and a half. (I: Would you say, that’s, um, are you using less frequently or are you using less each time you use?) Less each time. (I: Less each time. So about the same frequency throughout the day?) Right. But like I said, you can’t even do a full bag without being in like a, a zombie-like state.

Crack cocaine, actually. I’ll do maybe a bag [of dope] once a month, honestly. It’s like barely once a month, if that. Because I’m scared of the xylazine. If I can’t get regular, I will not do it. Absolutely not. (I: So your dope habit, like as far as opioids goes, is like pretty minimal right now?) Oh, yeah. If at all, honestly, because I’m actually on methadone.

Yeah, I was using less. (I: Okay. Was that, uh, less times during the day, or were you using less, like, when you would use? Would you use a smaller amount?) Yeah, smaller amounts. You only need like a half of a stamp bag because it’s so strong.

Yes. Decreased quite a bit. Um, because we were, we were, we were pretty, I mean, I consider it pretty heavy usage for us. Um, we’d buy, like, a brick a day. And then, once the tranq showed up, within, like, a month, we got down to just splitting a bundle a day. So, five bucks a piece as opposed to 25 a piece. Um, the other reason for that is that the tranq is strong. I mean, where it’s not the same high.

I used to do like a brick a day. Do you know how much that is? (I: Yeah. Yeah.) And then I got that down to like three buns a day. And then like now I do like at most a bun a day.

##### Recovery Aspirations

I don’t know, in a way I’m kind of trying to look at it as a blessing now, but I’m at the point, I’ve tried going through rehab several times, but I know this is the time, you know what I mean? Because I’m not even, I’m not even getting to you know, where you can function normally. You know, most people try to use, keep using so they can function. I’m not even getting that anymore. You know what I mean?

It’s nasty, yeah, nasty. Well, I mean, yeah, and it’s been getting to the point where, I mean, if everything is going to have so much xylazine in it, I mean, it’s making us want to just not use at all. I mean, like, there’s a lot that goes into that, obviously. I’ve been getting high for a long time. My boyfriend’s been getting high for even longer to the point where he’s really, he wants to be done for a lot of different reasons, and I’m getting there too, but this is just hurrying it on a lot more. It’s just not satisfying, it’s not fun. It’s disappointing and aggravating, you know. There’s a lot of negatives about it that make us feel like it’s not worth it anymore. And it has to be pretty worth it to want to live this lifestyle. It has to be pretty darn worth it, and if it’s not, then you’re kind of looking at it and wanting to be done. (I: Yeah, you may have a bunch of reasons you might want to quit using. But now you’re not even getting what you would expect out of it.) Right, yeah, yeah. Exactly, there’s a lot of things that you will accept for, if it was going to be as good as it was when you first started. You’ll go through a lot of bullshit to get there, but if it’s not even going to be that, then you’re kind of done. So, we’ll see.

Um, well my drug use, I started off like using drugs at a young age. I started with like marijuana, alcohol, and progressed to cocaine, ecstasy, stuff like that. And like, I got into my early 20s, I started messing with opiates, and that’s been an on-and-off thing constantly since then. It’s tough kicking. I mean, yeah, I mean with the tranq and stuff now, it’s making it, making me not want to do it anymore.

I don’t. Yeah, I don’t like them. I don’t like them. Probably be the reason I quit, you know?

##### Quitting

Yeah I keep telling myself, I’m going near a week now, you know, my use, I’m so excited for this. (I: Yeah, that’s exciting.) It is. But, and it’s weird because the xylazine is so new, but that’s part of the reason what’s getting me to the point where I’m finally like, fuck this, you know what I mean? Like it’s eating my skin. Like hell no, it ain’t worth it.

Yep, that was one of the reasons why I, one of my many reasons why I quit using to begin with is I, thanks to the drug checking program, I knew what was in my stuff and an approximation of how much and my main person I was getting stuff that was pretty much pure, it was more fentanyl than anything else. And my friends when I wouldn’t be able to go through my main guy, they were getting stuff that consistently had xylazine in it. And it’s no lie, you get abscesses from those, I got one on my leg somewhere, I didn’t use, and the scab on it lasted for three to four months. I’ve never had anything like it before. It was just, and that particular drug, just everything about it irritates me. I literally felt dumber after every time I did it. It was the tranquilizing effects totally, I felt like a zombie. But I’m very very much against it. I’m very grateful for you guys for being able to know, that and also I’ve used, I brought stuff in so I know exactly, but I’ve used an awful lot of the individual testers to test before you ingest. When you’re hooked on opiates, it definitely makes sense, it’s the best practice to do it. But when you’re sick you’ll end up, or dope sick or whatever, stuff like that kind of goes out the window. But it’s still, knowledge is power.

It’s nasty. Yeah. I mean, that’s one of the biggest reasons. I mean, other than wanting to be clean in general, that was, it might have been a bonus, honestly, in my case, because it scared me to the point where, like, I knew I had to get clean. Because, like, I mean, I have scars from it because it eat away your skin. Like, even when you don’t miss, it’ll eat away. It’s like, that’s nasty. I mean, thank goodness I was able to stop when I did because people are not getting reversed as easy and people are dying and it’s sick. It’s sick.

I noticed it first, actually, because I have, I was, like, a terrible drug addict because I have really bad veins in general. So, it would be hard to hit and I noticed it first because the first time I missed when it was in it, it was, like, gasoline. Just, like, in my skin. It was so bad. And, I mean, it’s, typically, it would be, if you missed, it would get itchy and, like, red and bumpy. But, like, you wouldn’t feel it as much as you missed other than a little discomfort. This one, as soon as you missed, it was, like, lightning under your skin. It just was terrible. But, like I said, in my case, I think it kind of helped me because it had me more scared than anything. And I had a good amount of clean time before using recently because I slipped during COVID. And that’s when I relapsed. But before that, I had almost 15 years clean. So, I really screwed up a good thing, but I have a greater appreciation for it now.

And the tranq probably, too. (I: Yeah, yeah.) That’s, that’s the, the thing that really, and that, I mean, it kind of is a, a good thing because that’s really why we stopped.

I haven’t been using dope in like 30 days. So, like 30 days ago. (I: Yeah. You said you use the xylazine test strips?) Yes. (I: What happens if it tests positive?) We throw it away. (I: Throw it away?) Yeah, like, for real. Nobody wants that shit where I’m from.

##### Quitting (MOUD)

Because, I don’t know, I guess the grams last me longer, you know what I mean? Typically a gram lasted me two or three days, and now it’s lasting me like four days. But now it’s not even doing anything like I can’t [get through the day?]. I don’t know if I’m addicted to the xylazine or the fentanyl or both or what it is. But yeah, like it’s not even making me right anymore. So I got back into the methadone clinic. (I: Gotcha, yeah. No, I mean I guess if it’s you know, it goes from like xylazine in fent to just xylazine, you’re not even getting –) Right, yeah. (I: -- an opioid anymore.) Nothing to even maintain, you know what I mean? So.

So it seems to me that I’m not getting the feeling of feeling better. I suffered a lot of trauma as a child. I have a lot of bad thoughts and memories that cloud my brain. Maybe that’s why I talk about shit all the time, so I don’t got to think about shit all the time. You know? Plus I just have a wealth of worthless knowledge. But you know? But I don’t get the gratification. I don’t get the medication that I’m looking for. And so now, I’m just like, like I do go to a clinic, so that stops me from being sick. So there’s no real drive for me anymore to… And another thing, I guess, is for a long time, at the clinics, they were kicking people out for using, which makes absolutely zero fucking sense to me, because my problem may be worse than your problem. You know? I’ve been in rehab probably 30 times. I quit counting at 28 times or some shit like that. I used to go once a year, you know? Because I wanted to quit. I never wanted to do this with my life. I just wanted to feel better. You know? … So I suffered trauma. You know what I’m saying? So I mean, I was self-medicating. But it’s not working anymore. Like, the medication that people are whipping up in their kitchen right now isn’t heroin. And it’s not even real opiate. You know, it’s a fucking tranquilizer. I don’t want to go to sleep. I want the chemicals in my brain to make me feel better. And it’s not happening. That’s the difference, I guess.

Um, so, so say I would like shoot up in this arm. Um, then this arm would get an abscess. Or like, I would shoot up in this leg and then this leg would get an abscess. And I was like, what is going on here? So that, that, that’s what changed. Um, I tried to cut back because, you know, it would, it would burn my skin essentially. Um, and that’s what made me get on Subs. But even on subs, like, I mean, these used to be a lot deeper. Where now, like, my skin’s at least lining up with my skin. (I: Gotcha. So you, so are you saying you switched to Subs? Or you, you got on Subs because of the xylazine?) Yeah.

##### Route of Administration

Uh, yeah. Yeah. Yeah, just because, um, either they blew all their veins. I’ve tried to smoke it. But at the time my tolerance was really high. So I don’t really get much of a feeling from it. And then I tried to go back to snorting. Cause I snorted dope for about, like, two and a half years first. So then I was always in the ER for my asthma. And then, um, I can’t hit myself. And so, I mean I can. But it’s like a lackluster, maybe I’m hitting this? Like, for a while, like, whenever I saw blood, I thought, like, it’s a hit. But really it’s just cause the needle was almost all the way out of my skin. But I have seen, like, um, I would say, like, six people that I knew used to shoot up have been, um, now smoking it. (I: Have you heard any other reasons people are switching?) The xylazine. Um. I don’t know if, um, you can still get the wounds if you snort. Um. Like, it’s kind of embarrassing to say, but, like, one time I tried to boof it, thinking, like, oh, maybe that would help. But, um, I don’t, like, I don’t know, I didn’t really want to stick anything on my ass. And I read online, like, you can, like, front boof it. So I thought, like, oh, it’s kind of like a tampon. But then after I did it, I was like, oh, my God, if this is what it did to my legs, what if it does it to my vagina?

Yeah, like a lot of people are like smoking this stuff now. Like, bags. I noticed that, like a lot of people ain’t shooting dope no more. Because it’s scary man, you know what I mean. Just like going to sleep anywhere, like, you can’t even wake somebody up.

I’ve noticed a lot of people, yes, like try to like, yeah, switch over to snorting or smoking it. Like I never knew hardly anyone that smoked it a couple of years ago. And now I know a lot of people. (I: Right. Right. Yeah. I’ve noticed that especially amongst younger people.) Yeah. Yeah. Younger kids. I’ve even snorted. I’ve even snorted sometimes when I just try to just, you know, even I can’t always shoot up all the time. (I: Right. Right. Yeah. Is that because you have trouble finding veins and shit?) Yeah. And also, like I try to like sometimes I don’t want to just be out. Nodding out. I like to be there.

Yeah. A lot of people are smoking it now. A lot of people started smoking it. I’ve even tried it. But, yeah, I don’t know. (I: Doesn’t do it?) My one buddy who I usually get from, he hasn’t had what I like, he only had like a hardcore trippy shit for a minute. And like I was sick for like a week. It was awful. I just like refused to do it. Going all around town talking to people. And it was like everyone had the same shit, the exact same bag. It fucking sucked. Yeah. It was awful. (I: So people are smoking.) Yeah.

I don’t know. Like, like I know a lot of people that never even used that are just like smoking now, which I’ve noticed. Fucking, they just like started doing it. A lot of people prefer it. Like prefer that shit. It’s crazy. But, uh. (I: So people that never used it before are starting with smoking.) Yeah. I noticed a lot of that, yeah. At least they’re smoking instead of just going straight to the needle. (I: Right. Anybody like snorting or sniffing?) Yeah. Yep. People, veteran people. Like, it’s also like been shutting down veins. Like, me and my best friend, fucking he’ll tell you. But, you know, we’ve been shooting up like two decades, you know. Like, no veins anymore. It’d be like fucking hours of just sitting there trying to hit. I don’t know what it does, but it’s just. I mean, look what it does to your fucking, IM it, when you miss. Like, what it does to your tissue. Like, imagine what it’s doing to the inside of your fucking veins. (I: Right. Yeah. I heard somebody say it felt like there’s gel in their veins.) Yeah. Like, jelly blood. That’s funny. I call it jelly blood, too. (I: You call it jelly blood?) Yeah. Jelly blood. It turns your shit into like maple syrup. (I: Really?) Like, literally jelly. It’s crazy. Yeah. So it ruins your shots. Like, once you get little blood in there, it just coagulates and you can’t even get it. Just waste it. People are just, like, smoking it. Or snorting.

Right, right. It’s like, well, honestly, ever since then, I started, well, not ever since then, I would say the past month, I started snorting it instead of shooting it. (I: You moved from shooting to snorting?) Yeah. (I: You said in the last month or so?) Yeah, yeah. Like, my boyfriend, he snorts it. And, like, he had to Narcan me and it was, like, ridiculous. Like, anywhere you go, I would be, like, I mean, it’s sad because he’s, like, recorded me and showed me it. And I’m, like, it’s embarrassing, you know what I mean? Like, it’s, like, everybody can tell you’re definitely on something. (I: Right. Has snorting helped at all?) Yeah, yeah. Yeah, it’s, I noticed, like, I just, like, yeah, I, I mean, it’s not, like, like, it used to be, but it’s still not as bad as, like, injecting it. (I: Yeah. What’s changed or what feels different when you snort it?) Um, well, it’s, it’s just, like, I probably would say I do get, like, a little bit of a nod where it’s, and it doesn’t, like, knock me out. I don’t know, as much, like.

Yeah, that’s the only thing that’s really around right now, honestly. (I: Yeah. Do you use dope?) Yes. I was clean for five years. I just started using again like a year ago. (I: Yeah. So was it around when you started using again?) Yeah, that’s like the only thing that’s really around right now, honestly. I used to be an IV users but I only sniff right now. But people that use IV, it’s just horrible. It’s like eating away their arms. Looks like beehives. I’ve seen a girl with actually maggots in her arm. Yeah. It’s tearing apart my nose. (I: Is it?) Oh my gosh, I have a hole from one side to the other. (I: And that’s since the xylazine?) Yup.

First of all, I shouldn’t be doing it at all. Not that I think that sniffing is better than IV, but the needle, I think I was more addicted to that too. I wanted to shoot up coke and everything, you know, it was nasty. You know what I mean? It made my arms look disgusting, you know what I mean? Like, I don’t know, I really just look like a total snap. You know what I mean? I don’t know. Not that any way is good or the, you know what I mean? But I don’t know. (I: Yeah, no, I’ve heard people say, yup. Addicted to the needle more than anything.) Right, yeah. You miss and then it makes you, things on your arms, I had abscess, I would get cellulitis and stuff. Like, it was bad. Like, not saying that like, I’m, you know what I mean, any better now, but I’m going to get into rehab actually.

I’ve been, I stopped injecting for a while, and I, this is embarrassing to say but, I just do anal, like I just do it, I put it up my butt. But sometimes I still try to hit, but it’s like, it leaves, it’s weird, it’s almost like a mess, what it’s doing to your body, so I just don’t even try it anymore. (I: Is that because of the xylazine, or just because you’ve been injecting a while?) I, no, I think it’s the xylazine, because everybody that I know is the same thing’s happening to them, so… (I: So people are switching away from injecting?) Well, I don’t know, the other people know, because I, a lot of people, I know their arms are about to fall off, and they got bumps all over their body, and… Yeah. You know, I don’t know, yeah, it’s not good. And also like it clogs up or something, I don’t know what it does, but it’s just, yeah. It’s not real, that’s for sure, it’s not real heroin.

##### Using Differently

Um… I… If I want… If I get the tranq, I like to do it just at night because it puts me to sleep. Um… But… I don’t like doing it during the day because it makes me lose my memory. I don’t… I’ll like wake up in places I don’t know where I’m at. Um… Or I don’t remember getting there. It makes… I guess it just makes you black out.

A while ago I was. Uh, I’d say probably, uh, maybe a year ago. I was, I remember everyone was like, I wish I could find something that didn’t have xylazine in it. But then everybody who found xylazine in it, um, like, as soon as, you know, anybody was like, I’m like, oh, this dope don’t have xylazine. Well, then they would do it and be like, oh my god, I feel sick now from, and I want the ones *with* xylazine. So, how I would do it would be like, one, one with xylazine and one without xylazine, you know? (I: Oh, okay. Take one of each?) Yeah, I would, like, alternate. (I: Gotcha. Okay. So, do you get, uh, sick if, if, if you don’t have xylazine? Like, do you get the same kind of?) Yeah, I think, I think, like, um, I know, I noticed, like, at the, I started going to the methadone clinic, and I noticed that the methadone clinic will raise you a lot higher because of the xylazine, and, and I noticed, like, before I wouldn’t, like, the methadone would hold me a lot longer.

#### Reasons for Changes in Use

##### Not Experiencing Intended Effects

So it seems to me that I’m not getting the feeling of feeling better. I suffered a lot of trauma as a child. I have a lot of bad thoughts and memories that cloud my brain. Maybe that’s why I talk about shit all the time, so I don’t got to think about shit all the time. You know? Plus I just have a wealth of worthless knowledge. But you know? But I don’t get the gratification. I don’t get the medication that I’m looking for.

Oh, yeah. Because, first of all, it’s, it’s less, like, heroin, when you do heroin, it feels good. Obviously. That’s why we do it. (I: You get the warm, the warm hug.) Yeah. But, that shit, it doesn’t. Like, it doesn’t, it’s not like, and then, obviously, it’s too late when you do it.

Well, I mean, I’ve been using, honestly, for like, since I was 20 and I’m 47. I’m off and on. I’ve had some clean time, but I know since, like, I remember before xylazine was involved, I remember, you know, like, the high, it was like, you know, you just nod. Well, and then I remember before I knew xylazine was in it, I remember thinking, this is weird, you just do it, you pass out, and then you wake up and need to do more. Like, there was, like, really not much of a nod, you know what I mean? It was just like, what’s the point of doing this?

Mm-hmm. Oh, yeah. And, and just the fact that, like, it’s not actually enjoyable. Like, you know, it doesn’t actually make you, it doesn’t give you the same feeling. So, it’s a lot easier to stop because of that.

I mean I guess if it’s you know, it goes from like xylazine in fent to just xylazine, you’re not even getting –) Right, yeah. (I: -- an opioid anymore.) Nothing to even maintain, you know what I mean? So.

##### Physical Health Risks

But, like, I, it made me hallucinate and shit before. Um, and, like, it’s just not, and then the biggest reason was my wife, um, got a couple little, of the little wounds on her. And I’m like. (I: They’re like chemical burns.) Yeah. (I: They take, like, six months to heal.) And, well, in my, at our clinic, there’s a lot of people that have had, had to have limbs removed and shit from it. And, um, I’m like, if, if something like that happens and she has to get her leg amputated or her arm amputated, like, cause it’s, I’m the one who fucked up. And, like, and she just followed me. So, I’m like, if, if I fucking, like, I feel like it would be my fault and I’d never fucking—It’s nasty. Yeah. I mean, that’s one of the biggest reasons. I mean, other than wanting to be clean in general, that was, it might have been a bonus, honestly, in my case, because it scared me to the point where, like, I knew I had to get clean. Because, like, I mean, I have scars from it because it eat away your skin. Like, even when you don’t miss, it’ll eat away. It’s like, that’s nasty.

I noticed it first, actually, because I have, I was, like, a terrible drug addict because I have really bad veins in general. So, it would be hard to hit and I noticed it first because the first time I missed when it was in it, it was, like, gasoline. Just, like, in my skin. It was so bad. And, I mean, it’s, typically, it would be, if you missed, it would get itchy and, like, red and bumpy. But, like, you wouldn’t feel it as much as you missed other than a little discomfort. This one, as soon as you missed, it was, like, lightning under your skin. It just was terrible. But, like I said, in my case, I think it kind of helped me because it had me more scared than anything. And I had a good amount of clean time before using recently because I slipped during COVID. And that’s when I relapsed. But before that, I had almost 15 years clean. So, I really screwed up a good thing, but I have a greater appreciation for it now.

So it’s like there’s no point, I’m getting straight xylazine a lot of times. But yeah, my legs are tore up dude. They’re, they’re horrible. They’re really bad. So I go to the wound clinic and everything because it’s yeah, just eats up your skin so yeah. But yeah, it’s it’s it’s an evil thing the xylazine, who knows what it’s doing inside. I’m actually going through a lot of medical stuff right now, just trying to figure out why my heart rate’s not where it should be, like all kinds of, like my kidneys, all kinds of stuff. And I think it’s because of the xylazine. It’s the only thing that’s been different, you know this past year than the 10 years before that, that my use would have had in it, you know what I mean?

##### Functional Impacts

I, I, like, I hate it so much. I miss a lot of appointments due to it because I will end up like, I’ll use it and then next thing you know, I’m waking up and my kids are walking in from school. I’m like, where did that time go? And I’m so confused. I’m just like, and I just can’t get my, I can’t wrap my head around it sometimes. I’m just like, where did this time go? And stuff, and I just, I, I don’t like it. (I: Yeah. Yeah. Yeah. I’ve heard a lot of people are using less, you know, either because, you know, the physical effects or because they can’t be, they can’t get anything done in a day.) Yeah. (I: It just knocks you out.) Yeah. And it’s, it’s ridiculous. I don’t know why they want to use xylazine. And it’s a horse tranquilizer, isn’t it? (I: Yeah.) Like, what was the point? What’s the point of putting it in there? I don’t understand. Yeah. And the sleepiness, that’s why, yeah.

The tranq, it’s like, you just fall asleep. Um, so, it’s, which, I’ve talked to some people who actually enjoy that, and I, whatever. But, um, personally, I mainly, what started my use back up in the beginning, and this time, is just my anxiety. So, I don’t really want to be sleeping. I just don’t want to be constantly freaking out. Which, in turn, when you start using, you give yourself more anxiety. So, it’s like, I ended up making it worse, but. Yeah. Yeah. It’s a vicious cycle.

Yeah, it’s like, you just go to sleep. And it’s like, I mean I don’t love this. (I: And you don’t really feel the euphoria.) No. (I: Like heroin used to be that warm hug feel.) Yeah, and then like, and now, like, you don’t know what the hell you’re doing. Like it’s just crazy.

Well, I use a lot less, that’s for sure. (I: Yeah, why’s that?) Just because you only need a little bit, if you do too much of it, you’re just going straight to sleep. You waste the whole day. You do two shots, one in the morning, wake up at two, three in the afternoon, do another one, waking up at midnight. It’s kind of dumb. (I: Yeah. So you use a little less, it just knocks you out.) Yeah, absolutely.

Uh, it decreased. Using a lot less. I’d probably go from a brick a day to probably about a bundle. To maybe a bundle and a half. (I: Would you say, that’s, um, are you using less frequently or are you using less each time you use?) Less each time. (I: Less each time. So about the same frequency throughout the day?) Right. But like I said, you can’t even do a full bag without being in like a, a zombie-like state.

##### Overdose Risks

It’s just like, it’s crazy. Like I’ve almost died off like one bag and I’m like a veteran with this shit. But like it doesn’t even make sense. You like do like put one bag in. It’s not even a big bag and you’ve got to do like a fraction of that and you’re fucking done. You know, like there’s some shit that like, like I hate like the trippy shit. It’s unreal. Like it’s, you know, I don’t even get well off of it. And it’s just like, I don’t know, it’s scary. You know, like I almost died off like the first, the first gnarly one. It was called Special Meds. And yeah, I almost died. I was like the first one out of everyone to do it.

It’s nasty. Yeah. I mean, that’s one of the biggest reasons. I mean, other than wanting to be clean in general, that was, it might have been a bonus, honestly, in my case, because it scared me to the point where, like, I knew I had to get clean. Because, like, I mean, I have scars from it because it eat away your skin. Like, even when you don’t miss, it’ll eat away. It’s like, that’s nasty. I mean, thank goodness I was able to stop when I did because people are not getting reversed as easy and people are dying and it’s sick. It’s sick.

Yeah, it made me a lot more concerned with things. I had started intentionally using less and less. I’ve done an awful lot of reversals and I know that particular one also affects your central nervous system and keeps you, Narcan doesn’t help reviving the respiratory part of it. So I had began using less and less until I had an intervention, essentially. I had a warrant I was running from for a while and it caught up to me, it was like two years ago. I went to a county jail and ended up detoxing in there. A whole lot of my overdose reversals have been from people from a period of abstinence or when they get out of jail, mostly when people get out of jail and go out. What is it? After two days, your tolerance is reduced by 50 percent. Just that alone. But the majority of the time, I couldn’t believe how little it would take for somebody to use again to go blue in the lips and quit breathing and need Narcan.

## References

Bedard, M. L., Huang, X.-P., Murray, J. G., Nowlan, A. C., Conley, S. Y., Mott, S. E., Loyack, S. J., Cline, C. A., Clodfelter, C. G., Dasgupta, N., Krumm, B., Roth, B. L., & McElligott, Z. A. (2024). Xylazine is an agonist at kappa opioid receptors and exhibits sex-specific responses to opioid antagonism. Addiction Neuroscience, 11, 100155. 10.1016/j.addicn.2024.100155

Cano, M., Daniulaityte, R., & Marsiglia, F. (2024). Xylazine in Overdose Deaths and Forensic Drug Reports in US States, 2019-2022. JAMA Network Open, 7(1), e2350630. 10.1001/jamanetworkopen.2023.50630

Carroll, J. J. (2024). Xylazine-Associated Wounds and Related Health Concerns Among People Who Use Drugs: Reports From Front-Line Health Workers in 7 US States. Substance Use & Addiction Journal, 45(2), 222–231. 10.1177/29767342231214472

Dasgupta, N., Brown, J. R., Nocera, M., Lazard, A., Slavova, S., & Freeman, P. R. (2022). Abuse-Deterrent Opioids: A Survey of Physician Beliefs, Behaviors, and Psychology. Pain and Therapy, 11(1), 133–151. 10.1007/s40122-021-00343-z

Dyer, O. (2024). Opioid crisis: Fall in US overdose deaths leaves experts scrambling for an explanation. BMJ, 386, q2091. 10.1136/bmj.q2091

Edinoff, A. N., Sall, S., Upshaw, W. C., Spillers, N. J., Vincik, L. Y., De Witt, A. S., Murnane, K. S., Kaye, A. M., & Kaye, A. D. (2024). Xylazine: A Drug Adulterant of Clinical Concern. Current Pain and Headache Reports, 28(5), 417–426. 10.1007/s11916-024-01211-z

Hays, H. L., Spiller, H. A., DeRienz, R. T., Rine, N. I., Guo, H.-T., Seidenfeld, M., Michaels, N. L., & Smith, G. A. (2024). Evaluation of the relationship of xylazine and fentanyl blood concentrations among fentanyl-associated fatalities. Clinical Toxicology (Philadelphia, Pa.), 62(1), 26–31. 10.1080/15563650.2024.2309326

Heidari, O., Sugarman, O. K., Winiker, A. K., Gattine, S., Flanagan, V., Razaghi, R., & Saloner, B. K. (2024). Personal Experiences With Xylazine and Behavior Change: A Qualitative Content Analysis of Reddit Posts. Journal of Addiction Medicine. 10.1097/ADM.0000000000001383

Hochheimer, M., Strickland, J. C., Rabinowitz, J. A., Ellis, J. D., Dunn, K. E., & Huhn, A. S. (2024). Knowledge, Preference, and Adverse Effects of Xylazine Among Adults in Substance Use Treatment. JAMA Network Open, 7(2), e240572. 10.1001/jamanetworkopen.2024.0572

Khatri, S. N., Sadek, S., Kendrick, P. T., Bondy, E. O., Hong, M., Pauss, S., Luo, D., Prisinzano, T. E., Dunn, K. E., Marusich, J. A., Beckmann, J. S., Hinds, T. D., & Gipson, C. D. (2024). Xylazine suppresses fentanyl consumption during self-administration and induces a unique sex-specific withdrawal syndrome that is not altered by naloxone in rats. Experimental and Clinical Psychopharmacology, 32(2), 150–157. 10.1037/pha0000670

Love, J. S., Levine, M., Aldy, K., Brent, J., Krotulski, A. J., Logan, B. K., Vargas-Torres, C., Walton, S. E., Wax, P., & Manini, A. F. (2023). Opioid Overdoses Involving Xylazine in Emergency Department Patients: A Multicenter Study. Clinical Toxicology (Philadelphia, Pa.), 61(3), 173–180. 10.1080/15563650.2022.2159427

Oyler, D. R., Slavova, S., Brown, J. R., Dasgupta, N., & Freeman, P. R. (2022). Kentucky pharmacists’ experiences in dispensing abuse-deterrent opioid analgesics. Journal of the American Pharmacists Association, 62(6), 1836–1842. 10.1016/j.japh.2022.07.017

Rodríguez, N., Vargas Vidot, J., Panelli, J., Colón, H., Ritchie, B., & Yamamura, Y. (2008). GC– MS confirmation of xylazine (Rompun), a veterinary sedative, in exchanged needles. Drug and Alcohol Dependence, 96(3), 290–293. 10.1016/j.drugalcdep.2008.03.005

Sadek, S. M., Khatri, S. N., Kipp, Z., Dunn, K. E., Beckmann, J. S., Stoops, W. W., Hinds, T. D., & Gipson, C. D. (2024). Impacts of xylazine on fentanyl demand, body weight, and acute withdrawal in rats: A comparison to lofexidine. Neuropharmacology, 245, 109816. 10.1016/j.neuropharm.2023.109816

Shover, C. L., Godvin, M. E., Appley, M., Pyfrom, E. M., Castrillo, F. M., Hochstatter, K., Nadel, T., Garg, N., Koncsol, A., Friedman, J. R., Molina, C. A., Romero, R., Harshberger, B., Spoliansky, J., Laurel, S., Jalayer, E., Ruelas, J., Gonzales, J., Snakeoil, S., … Sisco, E. (2024). Rapid emergence of UV stabilizer Bis(2,2,6,6-tetramethyl-4-piperidyl) sebacate (BTMPS) in the illicit fentanyl supply across the United States in July-August 2024: Results from drug and drug paraphernalia testing (p. 2024.09.13.24313643). medRxiv. 10.1101/2024.09.13.24313643

Sugarman, O. K., Shah, H., Whaley, S., McCourt, A., Saloner, B., & Bandara, S. (2024). A content analysis of legal policy responses to xylazine in the illicit drug supply in the United States. International Journal of Drug Policy, 129, 104472. 10.1016/j.drugpo.2024.104472

Wagner, K. D., Fiuty, P., Page, K., Tracy, E. C., Nocera, M., Miller, C. W., Tarhuni, L. J., & Dasgupta, N. (2023). Prevalence of fentanyl in methamphetamine and cocaine samples collected by community-based drug checking services. Drug and Alcohol Dependence, 252, 110985. 10.1016/j.drugalcdep.2023.110985

Watkins, D. C. (2017). Rapid and Rigorous Qualitative Data Analysis: The “RADaR” Technique for Applied Research. International Journal of Qualitative Methods, 16(1), 1609406917712131. 10.1177/1609406917712131

Wong, S. C., Curtis, J. A., & Wingert, W. E. (2008). Concurrent Detection of Heroin, Fentanyl, and Xylazine in Seven Drug-related Deaths Reported from the Philadelphia Medical Examiner’s Office. Journal of Forensic Sciences, 53(2), 495–498. 10.1111/j.1556-4029.2007.00648.x

